# *Vibrio cholerae* multifaceted adaptive strategies in response to bacteriophage predation in an endemic region of the Democratic Republic of the Congo

**DOI:** 10.1101/2021.07.30.21261389

**Authors:** Meer T. Alam, Carla Mavian, Marco Salemi, Massimiliano S. Tagliamonte, Taylor Paisie, Melanie N. Cash, Angus Angermeyer, Kimberley D. Seed, Andrew Camilli, Felicien Masanga Maisha, R. Kabangwa Kakongo Senga, J. Glenn Morris, Afsar Ali

**Author notes:** these authors contributed equally (Meer T. Alam, Carla Mavian). Corresponding authors: Carla Mavian, Ph.D., Department of Pathology, Immunology and Laboratory Medicine & Emerging Pathogens Institute, University of Florida, Gainesville, Florida 32601, Marco Salemi, Ph.D., Department of Pathology, Immunology and Laboratory, Medicine & Emerging Pathogens Institute, University of Florida, Gainesville, Florida 32601, Afsar Ali, Ph.D., Department of Environmental & Global Health & Emerging Pathogens Institute, University of Florida, Gainesville, Florida 32601.

## Abstract

Bacteriophage predation of toxigenic *Vibrio cholerae* O1 (the causative agent of cholera) has been linked with seasonal patterns of disease and with clinical response to infection in humans ^1-4^. However, we still lack a clear demonstration of how the interplay between bacteria and bacteriophage can influence shifts in strain populations. We analyzed toxigenic *V. cholerae* O1 isolated from patients in the Great Lakes, a cholera endemic region of the Democratic Republic of the Congo (DRC), between 2013-2017. Bayesian phylogeography shows that all strains derived from the East Africa T10 introduction event^5^, consistent with establishment of a regional endemic focus, and identified two major lineages, with the most recent correlating to ST515, a cholera cluster previously found in the Lake Kivu and expanding northward^6^. We also identified a novel ICP1 bacteriophage, genetically distinct from previous ICP1 isolates detected in Asia ^7,8^, from stool samples of cholera patients. The presence of phages in specific regions of the DRC resulted in the independent emergence, along both internal and external branches of the cholera phylogeny, of distinct mutational pathways in genes of the O1 biosynthetic gene cluster associated with phage resistance. Our data evidence, for the first time, *V. cholerae* multi-peaked adaptive landscape during outbreaks, and a complex co-evolutionary dynamic linked to presence of predatory phages.

---

Strains of the seventh cholera pandemic consisting of biotype El Tor strains (P7ET) were first reported in Africa in the early 1970s ^5^ and in the intervening years cholera has emerged as a major cause of illness in the African continent ^9-11^. The disease has been of particular concern in the DRC, with major outbreaks occurring in 2008, 2009, 2011-2012, 2013, and 2015-2017; in 2017 alone, an estimated 53,000 cholera cases with 1,145 deaths were reported from 20 out of 26 DRC provinces ^12^. Outbreaks have been most persistent in the eastern part of the DRC in the Great Lakes region, along the Albertine Rift ^11,12^. In work reported by Weill *et al* ^5^, strains appear to have been initially introduced into this area as part of what has been characterized as the T5 introduction under the “first wave” of the P7ET. In 1992, as part of the third wave of the P7ET, the disease was reintroduced by a strain from Asia, in what has been designated as the T10 introduction event ^5^. Subsequent studies have documented persistence of T10 strains in this region, and it has been suggested that the ongoing cholera outbreaks in the Great Lakes (and spread of strains from this region) reflect establishment of a regional focus of endemic disease derived from the T10 introduction ^6^.

As part of initial efforts to expand our understanding of *V. cholerae* O1 strain origins and evolution in the DRC Great Lakes region, we sequenced 24 toxigenic *V. cholerae* O1 strains obtained between 2015 and 2017 from DRC patients (Table S1). Strains were collected from stool specimens of cholera patients that attended cholera treatment centers (CTCs) from three provinces in the Great Lakes Region surrounding the city of Goma. Confirmation of the O1 antigen group was done by serologic testing. Twenty-one (87.5%) of 24 strains were *V. cholerae* O1 serotype Inaba, while 3 (12.5%) were serotype Ogawa, indicating that the two serotypes were co-circulating. All strains were *ctxB* genotype-I in wave 3 and were within the T10 introductory clade ^5,6^.

After confirming the presence of adequate phylogenetic and temporal signal (Fig. S1), we performed a Bayesian phylogeographic analysis to determine how the DRC isolates situated within a global context^13^, based on genome-wide high quality single polymorphisms (hqSNPs). The data set included the 24 DRC *V. cholerae* genomes obtained in this study, 71 publicly available genomes from outbreaks in eastern DRC between 2014 and 2016 ^6^, and 25 geo-temporally diverse T10 sub-lineage (and ancestor from India) toxigenic *V. cholerae* O1 isolates from across Africa and Asia ^5^ (Table S2). Consistently to the findings of Weill *et. al*. ^5^, our maximum credibility clade (MCC) tree (Fig. 1, Fig. S2) indicates that the introduction of T10 *V. cholerae* sub-lineage to Africa occurred between September 1991 - February 1996 (95% highest posterior density (HPD) estimate) with mean time of the most recent common ancestor (tMRCA) dating back to March 1994. The phylogeny suggests subsequent independent introductions (spillover events denoted by asterisk in Fig. 1) in the DRC Great Lakes region likely from Rwanda. The first one, in May 2001 (95% HPD September 1999 – June 2001), is represented by a DRC isolate, ERR1878097_CD_2003 (Fig.1), branching out of a lineage circulating in Rwanda. The other resulted in two major monophyletic clades that match previously reported MLST sequence types from Irenge *et. al*.^6^ (Table S3). The tMRCA of the first major lineage (denoted “*I*” in Fig. 1) dates to February 2009 (95% HPD November 2005 - September 2011) and corresponds to the ST69 cluster^6^ identified during the first reported outbreaks of cholera in the DRC during 2008 and 2009. However, the long branch separating the isolate from the Tanganyika province (ERR572559_CD_2013) at the basis of the monophyletic clade, suggests the possibility of unsampled *V. cholerae* strains (either from Rwanda or other neighboring countries) constituting a missing link between this DRC lineage and its actual ancestor (Fig. 1). The second major lineage (denoted by “*II*” in Fig. 1) contains all the Inaba serotype strains collected. According to the molecular clock calibration, lineage II tMRCA dates to September 1999 (95% HPD March 1999 - September 2000). The monophyletic clade also includes three strains, from Zambia and Niger, likely the result of spillover events from DRC, and further divides into two sub-lineages (“*IIa*” and “*IIb*” in Fig. 1) that diverged in November 2004 (95% HPD September 2002 - March 2007). Overall, molecular clock and phylogeography reconstruction (Fig. 1) suggest sub-epidemic circulation of cholera lineages in the DRC, moving northward in the eastern part of DRC, years earlier than the first reported cholera outbreaks in 2008-2009.

**Fig. 1.**
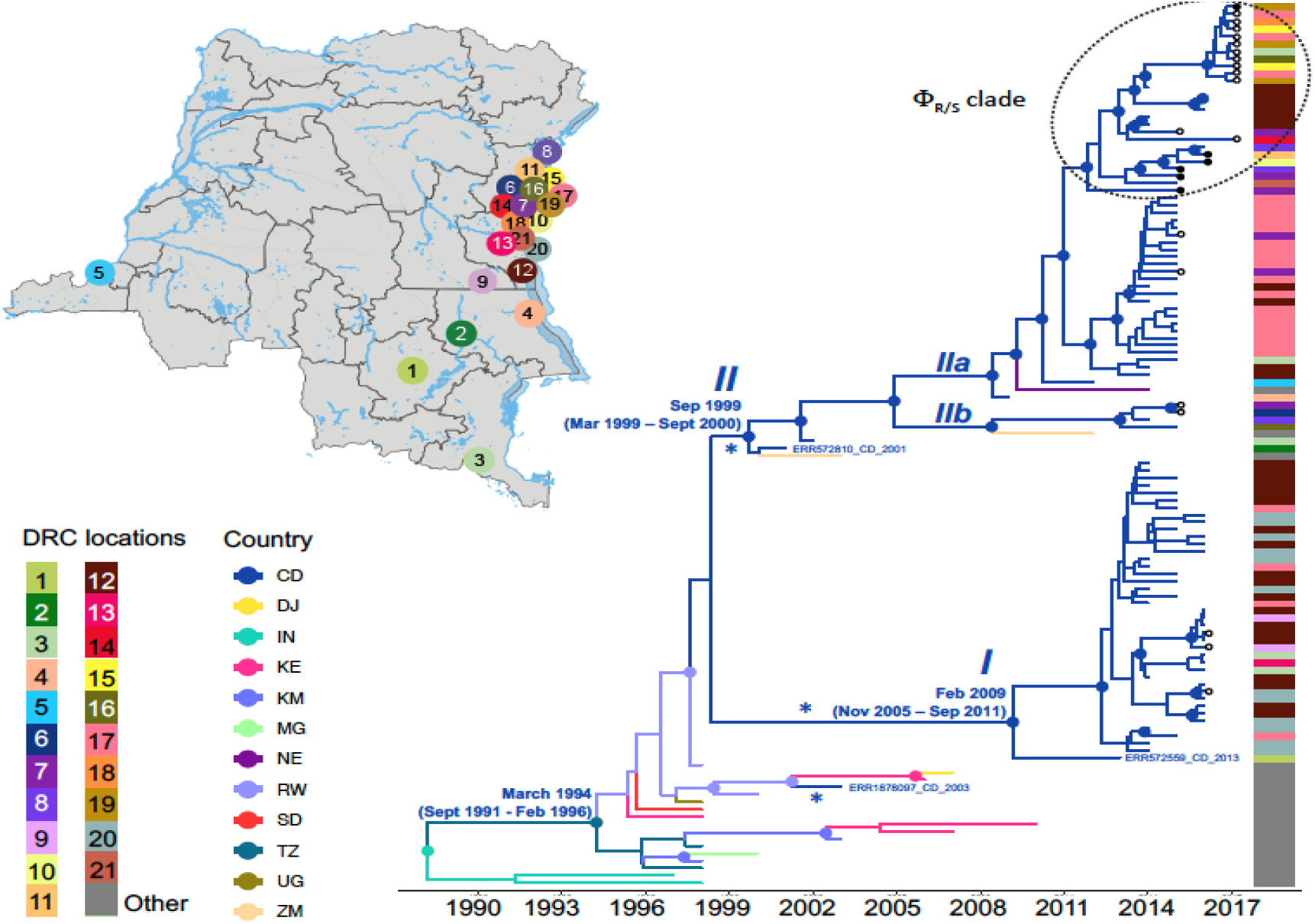
Spatiotemporal evolution and dissemination of *V. cholerae* DRC epidemic. The map indicates the sampling location of *V. cholerae* strains sequenced in this study. Each sampling location is coded by color and number. The locations are indicated for each tip in the Maximum clade credibility (MCC) tree as a heatmap (Table S2 for exact locations). The MCC tree was inferred from full genome *V. cholerae* isolates from DRC, neighboring African countries, and from Asia. Branches are scaled in time and colored by country of origin as shown in the legend (Country). The tree-associated heatmap indicates the location of the DRC samples in the map as shown in the legend (DRC locations), samples which do not have a DRC location are shown in gray. Circles in internal node indicate posterior probability (PP) support greater than 0.9 and the color indicate the ancestral country inferred by Bayesian phylogeography reconstruction. Circles at tips indicates the strains collected and sequenced in this study, with black circles designating phage resistant strains. Notations “*I*”, *“II”*, “*IIa*” and “*IIb*”, indicate DRC well supported lineages and sub-lineages circulating in the DRC during outbreaks. Asterisks indicate potential spillover events within the Great Lakes region originated from neighboring countries. The tree with full tip labels is provided in Fig. S2.

A separate collection of 41 stool samples obtained in 2016-2017 from cholera patients seen in CTCs in the Goma region (Fig. 2a) was screened for the presence of cholera-specific bacteriophages. Phages were identified and isolated from 17 of the 41 samples (Table S4), based on formation of clear plagues on strain AGC_15_CD_2017, one of the 24 toxigenic *V. cholerae* O1 strains sequenced as part of our phylogenetic studies (Table S1), which was randomly chosen for the phage plaque assay. Whole genome sequencing of a subset (n=8) of these phages showed that they were all highly clonal in nature (hqSNPs =114) and diverged substantially (hqSNPs = 8,441) from the ICP1 phage isolated from Bangladesh and India (Fig. 2b). Previously, a total of 185 core open reading frames were identified as being conserved in ICP1 isolates collected over a twelve-year period in Bangladesh and India ^7^. Uniquely, the DRC ICP1 lacks 15 of these core open reading frames (Fig. 2b), and also has 10.6 kb of novel sequence in the first third of the genome. Like the majority of ICP1-encoded gene products, most of the genes unique to DRC ICP1 are classified as hypothetical proteins due to a lack of an informative BLAST identification.

**Fig. 2.**
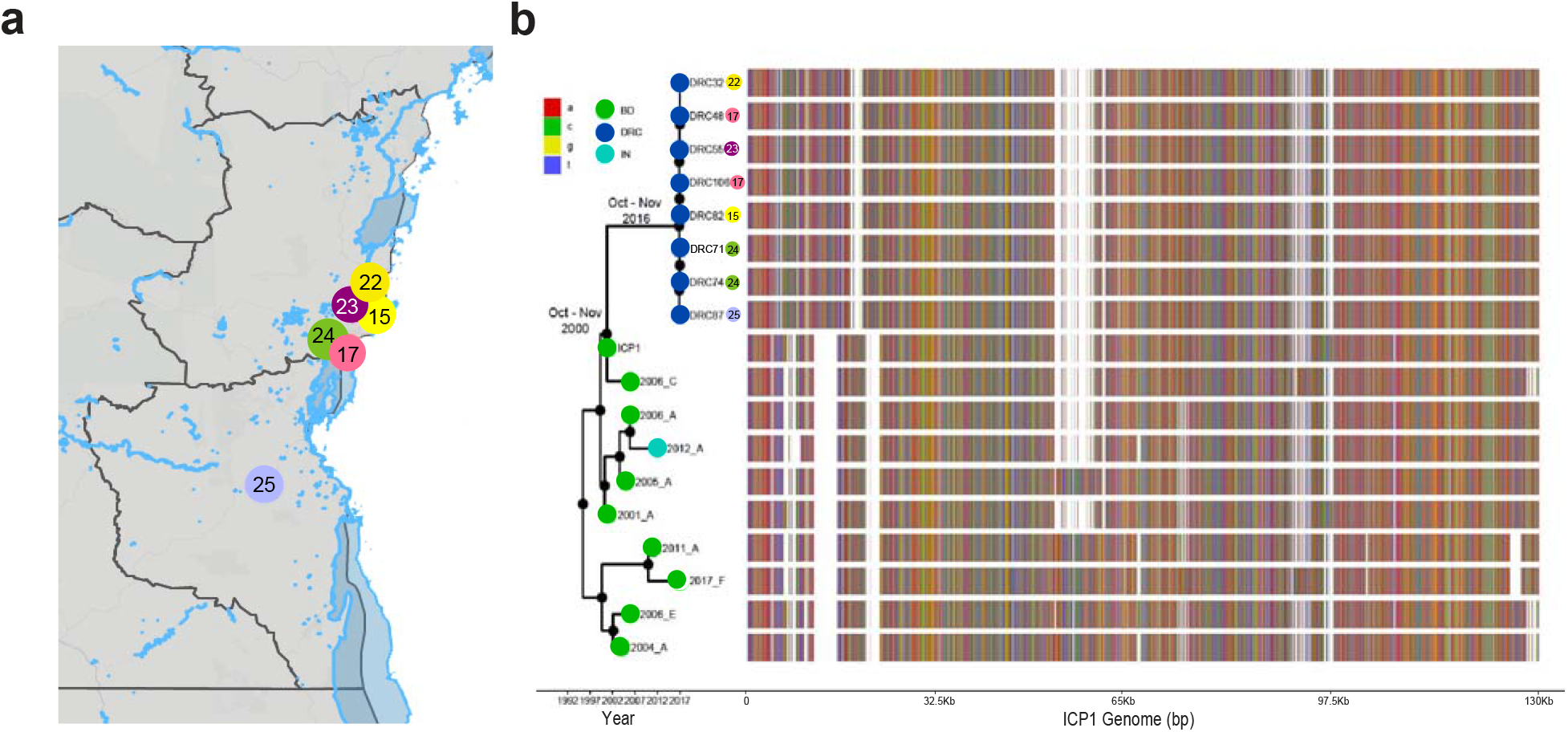
Bayesian inference of the phylogenetic relationship between DRC and Asian ICP1 phages and mutation patterns. (**a**) The map indicates the sampling location of ICP1 strains from DRC. Each sampling location is coded by color and number which also appear at the tip of the Maximum clade credibility (MCC) tree (Table S1 for locations). (**b**) The maximum clade credibility (MCC) tree branches are scaled in time and the circle tip points are colored by the location of origin: dark blue indicates isolates from the DRC, light blue signifies isolates from India, and green depicts strains isolated from Bangladesh. Circles in internal node indicate posterior probability (PP) support greater than 0.9. To the right of the MCC tree, the genomic composition of each isolate is displayed, with an adenine (a) indicated by a red marker, cytosine (c) is displayed by a green marker, guanine (g) is shown by a yellow marker, and thymine (t) is shown by a blue marker. White spaces indicate gaps at that location in the genome.

To investigate whether *V. cholerae* has been evolving in equilibrium with the newly identified ICP1 phages in the DRC, we determined the susceptibility of DRC *V. cholerae* O1 strains, isolated as part of the current study, to DRC ICP1 by plaque assays using ICP1_2017_A_DRC as reference. Of the 24 DRC *V. cholerae* strains, 18 (75 %) were susceptible to ICP1_2017_A_DRC, while the others displayed a phage-resistant phenotype (Table S1). At the genome level, resistant strains carried at least one mutation in genes that belong to the O1-antigen biosynthetic gene cluster (Table S5), which is not surprising as ICP1 uses the O1-antigen as its receptor, and *V. cholerae* undergoes phase variation to decrease and/or produce truncated O1-antigen production to evade ICP1 infection ^14^. On the other hand, other mutations usually associated with phage resistances, such as acquisition and expression of a family of phage-inducible chromosomal island-like elements (PLEs) ^15^, were not detected, and none of the DRC ICP1 isolates encoded, in fact, a CRISPR-cas system specifically targeting PLE for destruction, which would allow the phage to evade host immunity ^14,15^. However, our strains were still positive for O1-antigen by serology, indicating that O1-antigen is being produced, leading us to the hypothesis that low amounts of O1-antigen or a truncated version of that antigen can retain O1 antigen positivity by serology while inhibiting phage infection. Interestingly, comparison of genome-wide weighted averages of synonymous (*dS*) and non-synonymous (*dN*) substitution rates along the internal branches of the cholera phylogeny (Fig. 1) shows a dN/dS ratio significantly (p<0.001) greater than 1 (Fig. S3a). Moreover, the difference between *dN* and *dS* divergence accumulating over time along the internal branches of the phylogeny also appears to be increasing (Fig. S3b). In other words, the mixed presence of susceptible and resistant cholera phenotypes, at least in the DRC Goma region where samples were collected, together with dN/dS patterns suggest that *V. cholerae* has been evolving under pressure of increasing diversifying selection, possibly driven by the co-circulation of predatory phages. Indeed, the map of sampling locations shows that phage resistant or susceptible cholera strains, as well as independently sampled phages, tend to co-circulate in the DRC Goma region and surrounding locales (Fig. 3). At the same time, from a phylogeny perspective, all phage resistant cholera strains identified in our study appear to be intermixed with phage susceptible ones within a well-supported monophyletic clade of sub-lineage IIa (clade ΦR/S in Fig. 1).

**Fig. 3.**
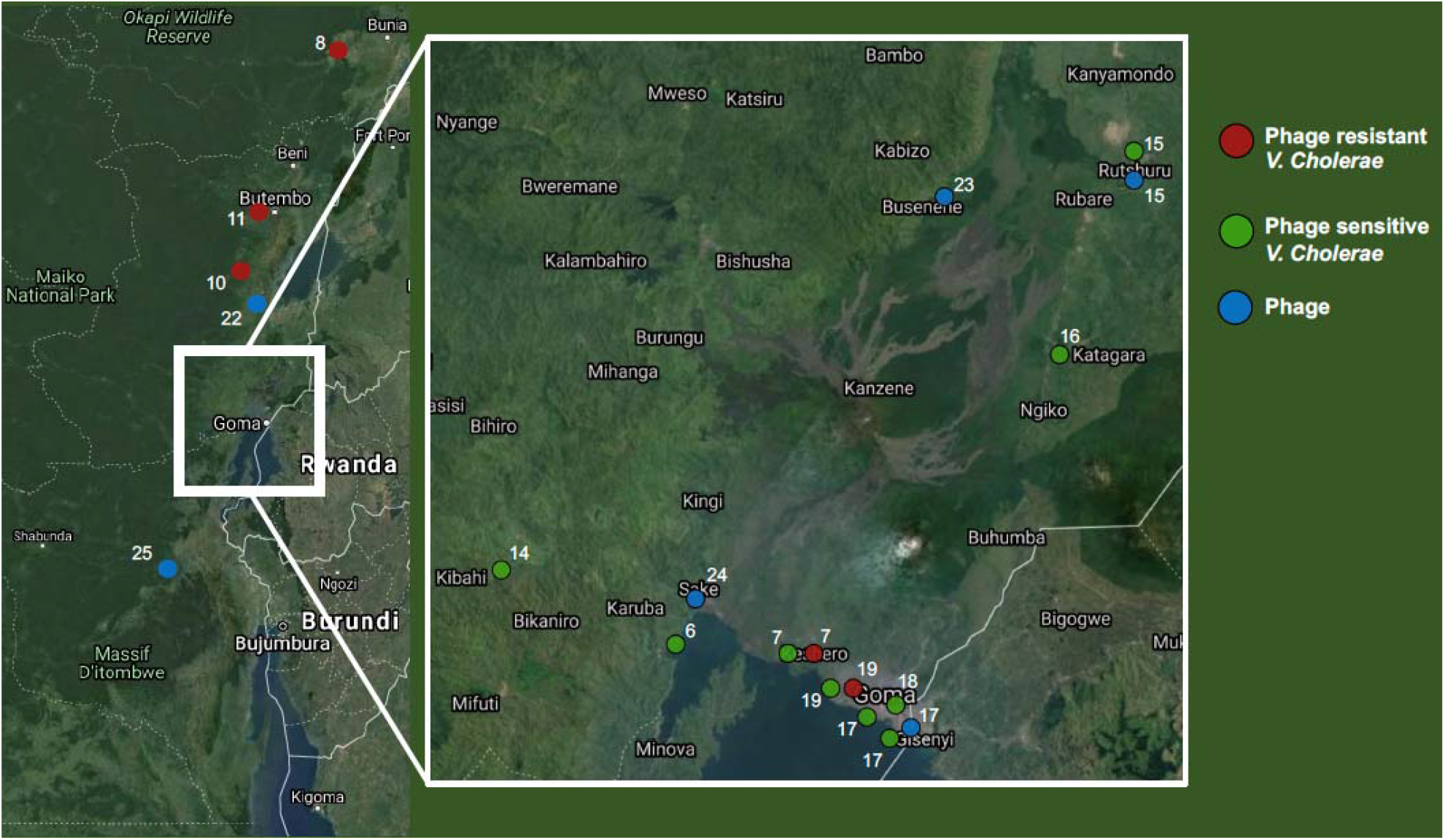
Sampling locations of phages and phage-resistant or sensitive f *V. cholerae* isolates in the DRC. The map indicates the sampling location of *V. cholerae* strains and the phages sequenced in this study. Each sampling location is coded by color, according to the legend to the right, and number which also appear at the tip of the Maximum clade credibility (MCC) tree for comparison (Exact coordinates of each location are provided in Table S2).

To examine in more detail the adaptive fitness landscape that would confer either resistance or sensitivity to phage predation, we optimized an MCC tree for the subset of cholera sequences including all strains in the Φ_R/S_ clade, as well as two outgroup strains (AGC-2-CD-2015, AGC-8-CD-2015) clustering outside the clade (Fig. 4). We used a Bayesian phylogeography model with phage resistance or susceptibility as discrete phenotypic characters (see Methods), to infer the most likely phenotype of the ancestral (internal) nodes of the tree. The analysis clearly shows that the backbone path (trunk) of the Φ_R/S_ clade, which represents the surviving lineage successfully propagating through time ^16^, connects phage sensitive ancestral sequences that generated, first, a sub-cluster of strains circulating in 2015-2016 and, then, a sub-cluster including 2017 strains (Fig. 4). In our data set, mutations in genes belonging to the O1-antigen biosynthetic gene cluster appear to have emerged, independently, along three distinct evolutionary lineages. The first one, leading to strain AGC-6-2015-DRC sampled in 2015, is characterized by amino acid substitutions in *RfbB V, rpIE, phrA, fliD* genes. The second one resulted in a monophyletic clade of phage resistant strains with mutations in either *rfbN* (strains sampled in 2015) or *rfbD* (sampled in 2016). The third one, leading to strain AGC-23-2017-DRC (sampled in 2017), characterized again by an amino acid substitution in the *RfbB* gene. Experiments using targeted mutagenesis in a host model mimicking human cholera’s pathology have shown a complex fitness landscape of *V. cholerae*^17^, including genes encoding for pathogenicity factors and growth. Yet, the parallel emergence of amino acid substitutions during outbreaks in different genes, each one potentially linked to phage resistance, is the first evidence within an epidemiology context of *V. cholerae*’s ability to explore a multi-peaked fitness landscape where adaptation to a new, hostile environmental condition (i.e., presence of predatory phages) can be achieved through different mutational pathways. It is worth emphasizing that every branch leading to phage resistant phenotypes in the phylogeny eventually dies out (Fig. 4), again underlying the multifaceted ability of cholera to explore and quickly abandon different evolutionary pathways during epidemic

**Fig. 4.**
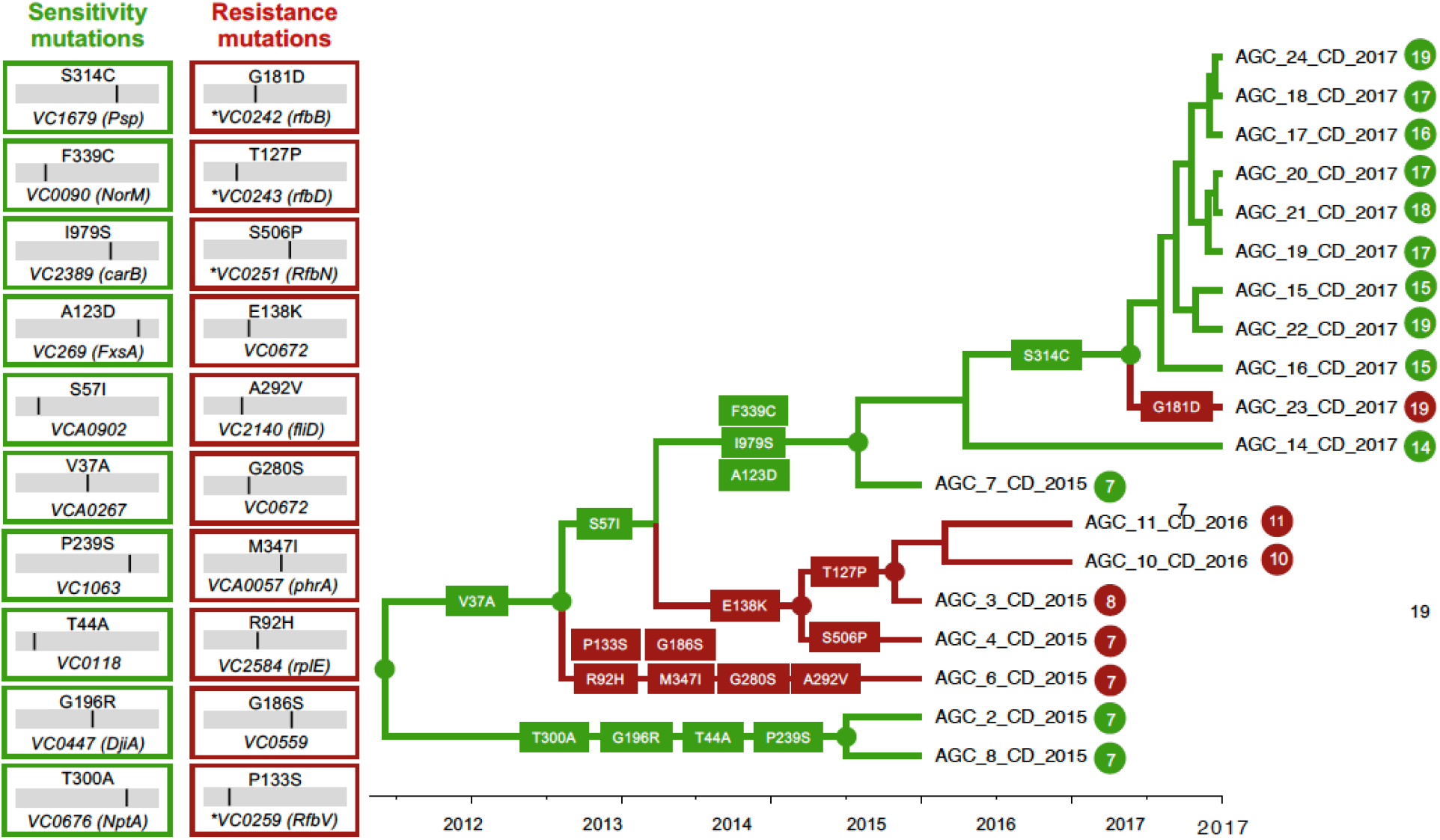
Phage-resistant or sensitive dynamics and mutational patterns of *V. cholerae* isolates in the DRC. The boxes show the mutations that have been found along the backbone, internal or external branches of the maximum clade credibility (MCC) tree. Each number is the amino acid position of the protein where the mutation was mapped in the tree. The name of each protein is displayed in the box to the left (as well as on the MCC tree) where an asterisk indicates a gene part of the O1-antigen biosynthetic gene cluster. The branches are scaled in time and colored based on resistance (red) or sensitivity (green). Circles and colors in internal node indicate posterior probability (PP) support greater than 0.9 for an ancestor to be resistant or sensitive.

spread. This is not surprising considering the fitness cost of phage resistance, resulting in mutants highly attenuated for virulence^18^. Of course, we are aware that one of the limitations of the present work is the small number of sites where either cholera strains or phages were collected, warranting caution in interpretating the data. Moreover, while we detected several mutations potentially linked with reduced density of O1-antigen such that ICP1 cannot fully engage the receptor to initiate infection, some mutations firmly associated by previous studies to phage resistance (e.g., frameshift mutations in *wbeL*) were not detected, while others (frameshift mutations in both poly-A tracts in *manA*) were found in the DRC cholera isolates but did not confer complete resistance to ICP1 (produced turbid plaques, see Table S5), as had been previously reported ^19^. Further work will be needed to determine the exact mechanism by which the *V. cholerae* strains isolated in the present study are either complete or partially resistant (turbid plaque) to ICP1_2017_A_DRC, and whether the phage can mutate to regain virulence as reported in earlier studies ^4^. Regardless of the mechanism, however, our findings supports a scenario where resistance to ICP1 in a subset of the *V. cholerae* strains contributes to the dynamic equilibrium of this predator-prey pair in the DRC.

In summary, our study suggests a complex co-evolutionary dynamic, involving *V. cholerae* and predatory phages, where phage sensitive and highly infectious strains co-circulate with phage resistant ones that occasionally emerge, and eventually die out, along different evolutionary pathways in response to predatory phages presence or absence in the environment, while the main phage sensitive evolutionary lineage keeps propagating through time. The ability of cholera to explore multiple mutational pathways in different genes and achieve phage resistance provides a significant evolutionary advantage in terms of quick adaptive response to a changing environment. Continuous monitoring of toxigenic *V. cholerae* and predator ICP1 phage in both patient stools and aquatic environments in the DRC, and elsewhere, could provide invaluable epidemiological data for monitoring the spread of cholera, understanding environmental actors driving successful dissemination, and assessing the potential for new outbreaks.

## Materials and Methods

### Isolation and characterization of toxigenic *V. cholerae* O1 and virulent phages

Between 2015 and 2017, stool samples from suspected cholera patients admitted in different cholera treatment centers around Goma, DRC, (Table S1) were placed on Carry blair transport media and brought to the Laboratoire Provincial de Sante Publique du Nord-Kivu in Goma for microbiological and serological analysis. *V. cholerae* in stool samples were enriched in alkaline peptone water as described previously ^20^. Following enrichment, a loopful of culture was streaked onto thiosulfate citrate bile salts (TCBS) agar and the plates were incubated overnight at 37°C. Bacterial colonies grown as yellow color on TCBS agar were subcultured onto Luria-Bertani Miller agar (LB-agar) and the culture plates were incubated overnight at 37°C. To determine serogroup, translucent colonies grown on LB-agar were tested against polyvalent antiserum specific for *V. cholerae* O1 and O139 by slide agglutination tests; each O1 positive strain was further typed for serotype using antiserum specific for Ogawa, Inaba or Hikojima serotype. The toxigenic *V. cholerae* were stored in soft LB-agar (0.7% agar) and sent to the Emerging Pathogens Institute at University of Florida (EPI) for further analysis. For isolation of potential virulent phages, the suspected cholera rice-water stool samples collected between 2016 and 2017, were centrifuged at 5,000 x g for 10 min in a microfuge. Stool samples used to detect and characterize virulent phages were different than the stool samples used for detection of 24 *V. cholerae* O1 strains described above. The resultant supernatant was filtered through a 0.22 µm syringe filter, stored at 4°C in a sterile microfuge tube, and sent to EPI for further analysis. We imported 24 toxigenic *V. cholerae* O1 strains to EPI using a global import permit issued by the USA Centers for Disease Control (CDC). At EPI, we again confirmed the identity of each of these isolates using standard microbiologic, serologic and genetic (PCR) screening tests as described previously ^16^. For detection and characterization of potential phages in cholera-confirmed patients’ stool samples, 41 stool sample filtrates were brought to EPI (Table S4).

### Virulent phage plaque assay

For detection and characterization of potential virulent phages, each filtered stool sample was tested by plaque assay using a host toxigenic *V. cholera*e O1 Inaba strain, AGC-15 (Table S1, S4). Isolated in DRC, AGC-15 has a wild-type *ompU* sequence, which encodes the receptor for ICP2 ^4^. AGC-15 also has the O1-antigen receptor for ICP1 and ICP3 ^7^ and lacks any PLE elements mediating immunity to ICP1 ^15^. Briefly, a sterile glass tube was inoculated with 100 µl of filtered-sterilized stool sample (potential source of virulent phage), 9.8 ml of LB-broth, and 100 µl of host *V. cholerae* AGC-15 culture (freshly grown to mid-exponential phase). The culture mixture was incubated overnight at 37°C with aeration to enrich any phages capable of infecting AGC-15. Following incubation, the culture was transferred to a 15 ml conical tube and centrifuged for 10 min at 5,000 x g at 4°C. To eliminate residual bacterial cells, the supernatant was filtered through a 0.22 µm syringe filter and the filtrate was stored at 4°C. The filtered supernatant was serially diluted (10-fold) in LB-broth to reach a dilution of 10^−5^ in a 96-well microtiter plate. One hundred µl of AGC-15 culture (grown to a mid-exponential phase) were added to the undiluted and each serially diluted filtrate in the microtiter plate and was incubated at room temperature for 10 min to allow phage adsorption. All 190 µl of each mixture from the microtiter plate was transferred to wells of six-well tissue culture plates. Three ml of soft LB-agar (0.35% agar) kept at 55°C in a water bath were added to each well of the six-well plate and the plate was gently swirled to mix the bacteria and phage evenly. To allow the agar to solidify, the plate was left for 30 min at room temperature followed by incubation at 37°C for 3-4 hours. After incubation, the plate was visually observed for plaque formation. If no plaques were observed after 4 hours of incubation, the plate was incubated overnight at room temperature to determine if any plaques were formed following extended incubation time. For virulent phage purification, a single clear plaque was picked using a Pasteur pipette into 1 ml of LB-broth and incubated overnight at 4°C to allow phage to diffuse out of the soft agar piece. High titer stocks of purified plaques were made by multiplication in broth culture as described above.

### Whole genome mapping and hqSNP calling

Toxigenic *V. cholerae* O1 samples collected were confirmed by serology and PCR ^20^. After subculture, gDNA extraction was carried out using the Qiagen DNeasy Blood and Tissue kit (Qiagen, Inc.). Genomic DNA from all isolates were cultured and extracted from a bacterial pellet. Sample library construction was performed using the Nextera XT DNA Library Preparation Kit (Illumina, San Diego, CA 92122). Whole-genome sequencing on all isolates was carried out with the Illumina MiSeq for 500 cycles (Illumina, San Diego, CA 92122). Adapter and raw sequence reads were filtered by length and quality by using the program Trimmomatic ^21^. After quality filtering, Bowtie2 ^22^ was used to map the sequence reads to the reference genome, *V. cholerae* O1 str. N16961 (GenBank Accessions: NC_002505.1 and NC_002506.1)^23^. After reads were mapped to the reference genome, duplicate reads were marked and realigned using Picard (http://broadinstitute.github.io/picard/). The reference-based mapping alignment was then verified and fixed accordingly. Freebayes ^24^ was used to create a custom genome-wide SNP calling database (dbSNP) from all isolates in the data set in order to perform base quality score recalibration (BQSR), as outlined in GATK’s best practice guidelines for germline variation (https://software.broadinstitute.org/gatk/). The newly created variant call format (VCF) file obtained from Freebayes ^24^ was subsequently filtered only for SNPs. The VCF file was used as a dbSNP for BQSR of the reference-based mapped alignment files; alignment files were then recalibrated and variants calling on the newly recalibrated files carried out with Freebayes ^24^. The newly created VCF file was filtered only for SNPs and normalized using the program BCFtools (http://www.htslib.org/doc/bcftools.html). Normalization simplifies the represented variants in the VCF file by showing as few bases as possible at particular SNP sites in the genome. SNPs were filtered by depth of coverage, quality, and genotype likelihood, as described in Azarian *et al*. ^25^. Finally, the SNP FASTA alignment was extracted by a custom python script from the VCF file. The SNP alignment was filtered site-by-site, leaving sites with only greater than 75% of SNPs at that particular site, making a high-quality SNP (hqSNP) alignment. Our hqSNP alignment then annotated using the program SnpEff ^26^. The final genome-wide SNP alignment included 120 T10 sublineage ^5^ *V. cholerae* genome sequences: 24 strains were collected in DRC (eight collected in 2015, five in 2016, 11 in 2017) and sequenced in this study; 71 were publicly available genomes from outbreaks in eastern DRC between 2014 and 2016 ^6^; six archival and publicly available DRC genomes collected between 2001-2013 ^5^; 17 genomes collected across Africa between 1998 and 2014 ^5^; and two publicly available genomes from India, ancestor of T10 sublineage ^5^. While the previously available DRC samples spanned 2014-2016 ^6^, we expanded the temporal dimension of the DRC collection as we sequenced mainly strains collected in 2017. The MLST analysis was performed on the online tool PubMLST ^27^ (Table S3).

### Phylogenetic and temporal signal

All datasets used in this study passed phylogenetic quality checks such as evaluating the presence of phylogenetic signal, to resolve the phylogenetic relationship among the *V. cholerae* isolates, and temporal signal for a robust calibration of the molecular clock (Supplementary Fig. S1). We performed likelihood mapping analysis using IQ-TREE ^28^, which allows the report likelihood values of the three possible unrooted trees, inferred using the best-fitting nucleotide substitution model, of each possible quartet (set of four sequences) on an equilateral triangle (likelihood map). In a likelihood map, dots (likelihood values) in the center of the triangle represent phylogenetic noise and simulation have shown that data sets with <35% nose (as it was in our case) can reliably be used for phylogeny inference ^29^. For each dataset, the presence of temporal signal was assessed by calculating the tree root-to-tip divergence regression plot with TempEst v1.5 (http://tree.bio.ed.ac.uk/software/tempest/)^30^, using maximum-likelihood (ML) phylogenies inferred with IQ-TREE ^28^ and the best-fitting nucleotide substitution model according to Bayesian Information Criterion (BIC) ^28^, and ultrafast bootstrap (BB) approximation (1000 replicates) to assess robustness of the phylogeny internal branches ^28^.

### Bayesian Phylogeography of DRC isolates

In order to test the hypothesis of whether cholera outbreaks in the DRC were caused by endemic *V. cholerae* O1 strains, or strains recently introduced from other African countries surrounding the Great Lakes region, we used the Bayesian phylogeographic ^13^ coalescent-based method ^31^ implemented in the BEAST v1.10.4 ^32^ software package. The reconstruction of *V. cholerae* O1 spatiotemporal spread from different locations through Bayesian phylogeography requires the calibration of a molecular clock. Evolutionary rates were estimated implementing a HKY nucleotide substitution model ^33^ with empirical base frequencies, gamma distribution of site-specific rate heterogeneity, and ascertainment bias correction ^34^, testing a constant demographic prior against non-parametric demographic models – Gaussian Markov randomfield Skyride (BSR) ^35^ and Bayesian Skyline Plot (BSP) ^36^ – to rule out spurious changes in effective population size inferred by a non-parametric model that would in turn impact timing of divergence events ^37^. Additionally, for each demographic model, we compared a strict and relaxed uncorrelated (lognormal distribution among branches) molecular clock ^38^. The best fitting molecular clock and demographic model were chosen by estimating the marginal likelihood of each model by using path sampling (PS) and stepping-stone (SS) methods, followed by Bayes Factor comparison test ^32,39^. A Markov Chain Monte Carlo (MCMC) sampler was run for 500 million generations, sampling every 50,000 generations. Proper mixing of the Markov chain was evaluated by the effective population size (ESS) of each parameter estimate under a specific model. ESS values > 200 for all parameter estimates are considered as evidence of proper mixing in the analysis. The sampling location for each isolate was used as a discreet trait in order to reconstruct likely locations of ancestral sequences (internal nodes in the tree) and infer migration events (bacterial flow) that took place in the DRC and throughout the Great Lakes Region. Phylogeographic analysis was carried out with the BEAST package v1.10.4 ^32^. Transitions between discrete states (location of where the isolate was collected) were estimated using the continuous-time Markov chain model operating the asymmetric migration model with Bayesian Stochastic Search Variable Selection ^13^. In our reconstruction of ancestral states, we assume migration occurs along branches connecting the tree nodes. The maximum clade credibility (MCC) tree chosen from the posterior distribution of trees using TreeAnnotator v1.10.4 after 10% burn-in. The MCC tree was annotated in R using the package ggtree ^39^ for publishing purposes.

### Calculation of weighted average of nonsynonmous (*dN*) and synonymous substitution rates (*dS*), and selection analysis

A codon alignment from the DRC clade was generated in order to analyze other mutations in the *V. cholerae* genome from the isolates in the phylogeny, and a subset of 200 Bayesian MCC genealogies randomly was obtained from the posterior distribution of trees for each subsampled data set. The weighted average of synonymous substitution rates (*dS*) and non-synonymous substitution rates (*dN*) in the protein-coding regions of the *V. cholerae* O1 genome for all, internal and external branches were obtained from a subset of 200 Bayesian MCC trees randomly obtained from the posterior distribution of trees, as described by Lemey et al.^40^, and also implemented previously ^16^. The subset of trees and in-house java scripts were then used to calculate *dNdS* rates and divergence in the isolates located within the DRC clade and plotted using the package ggplot2 in R.

### Whole genome sequencing, genome assembly and annotation of DRC phages

To characterize DRC phages, we plaque-purified phages from eight different patient samples, and prepared high titer stocks of each of these phages. Sample library construction for whole-genome sequencing was performed using the Nextera XT DNA Library Preparation Kit (Illumina, Inc.). Whole-genome sequencing was carried out with the Illumina MiSeq for 50 cycles (Illumina, Inc.). We obtained over 200-fold coverage facilitating de novo assembly of each phage genome into one complete contig using CLC Genomics Workbench (Qiagen, Inc.). Manual confirmation/correction of low coverage areas and/or problem areas were performed to ensure authentic genome assembly. Annotation of phage genomes was performed as described previously ^8^. Briefly, open reading frames from existing annotated ICP1 phages were compared against the DRC phage genomes using BLASTn to find homologues. Additional new putative open reading frames were discovered with de novo prediction software and added to the annotation. This was performed for each of the eight phages and due to very high similarity between them, one representative was randomly chosen and designated as ICP1_2017_A_DRC. This representative was aligned with existing ICP1 phage genomes using Mauve ^41^ and a maximum-likelihood, bootstrapped phylogenetic tree was generated with PhyML (-s BEST 92 --rand_start --n_rand_starts 10 -b 100) ^42^.

### Additional data

The genomic sequences have been deposited with NCBI under Sequence Read Archive (SRA) under BioProject ID: PRJNA748018. All other data are available in the Supplementary Information. R scripts and xml files are available upon request.

## Notes

### Competing Interest Statement

The authors have declared no competing interest.

### Funding Statement

Funded in part by NIH grant R01AI138554, awarded to JGM

### Author Declarations

UF Institutional Review Board

