## Supplementary figures and tables for "*Vibrio cholerae* multifaceted adaptive strategies in response to bacteriophage predation in an endemic region of the Democratic Republic of the Congo"

**Affiliations:** 1. Emerging Pathogens Institute, University of Florida, Gainesville, FL, USA; 2. Department of Pathology, Immunology, and Laboratory Medicine, College of Medicine, University of Florida, Gainesville, FL, USA; 3. Department of Plant and Microbial Biology, University of California, Berkeley, Berkeley, CA, USA; 4. Chan Zuckerberg Biohub, San Francisco, CA, USA; 5. Department of Molecular Biology & Microbiology, Tufts University, School of Medicine, Boston, MA, USA; 6. AMI-LABO, Goma, North Kivu Province, Democratic Republic of the Congo; 7. University of Goma, Department of Clinical Biology, Goma, North Kivu Province, Democratic Republic of the Congo; 8. Department of Medicine, College of Medicine, University of Florida, Gainesville, FL, USA; 9. Department of Environmental and Global Health, College of Public Health and Health Professions, University of Florida, Gainesville, FL, USA

#= these authors contributed equally.

*Corresponding authors:

| Carla Mavian, Ph.D.  Department of Pathology, Immunology and Laboratory Medicine &  Emerging Pathogens Institute  University of Florida  Gainesville, Florida 32601  cmavian@ ufl.edu | Marco Salemi, Ph.D.  Department of Pathology, Immunology and Laboratory Medicine &  Emerging Pathogens Institute  University of Florida  Gainesville, Florida 32601  | Afsar Ali, Ph.D.  Department of Environmental & Global Health &  Emerging Pathogens Institute  University of Florida  Gainesville, Florida 32601  |
| --- | --- | --- |

**Supplementary Figures**

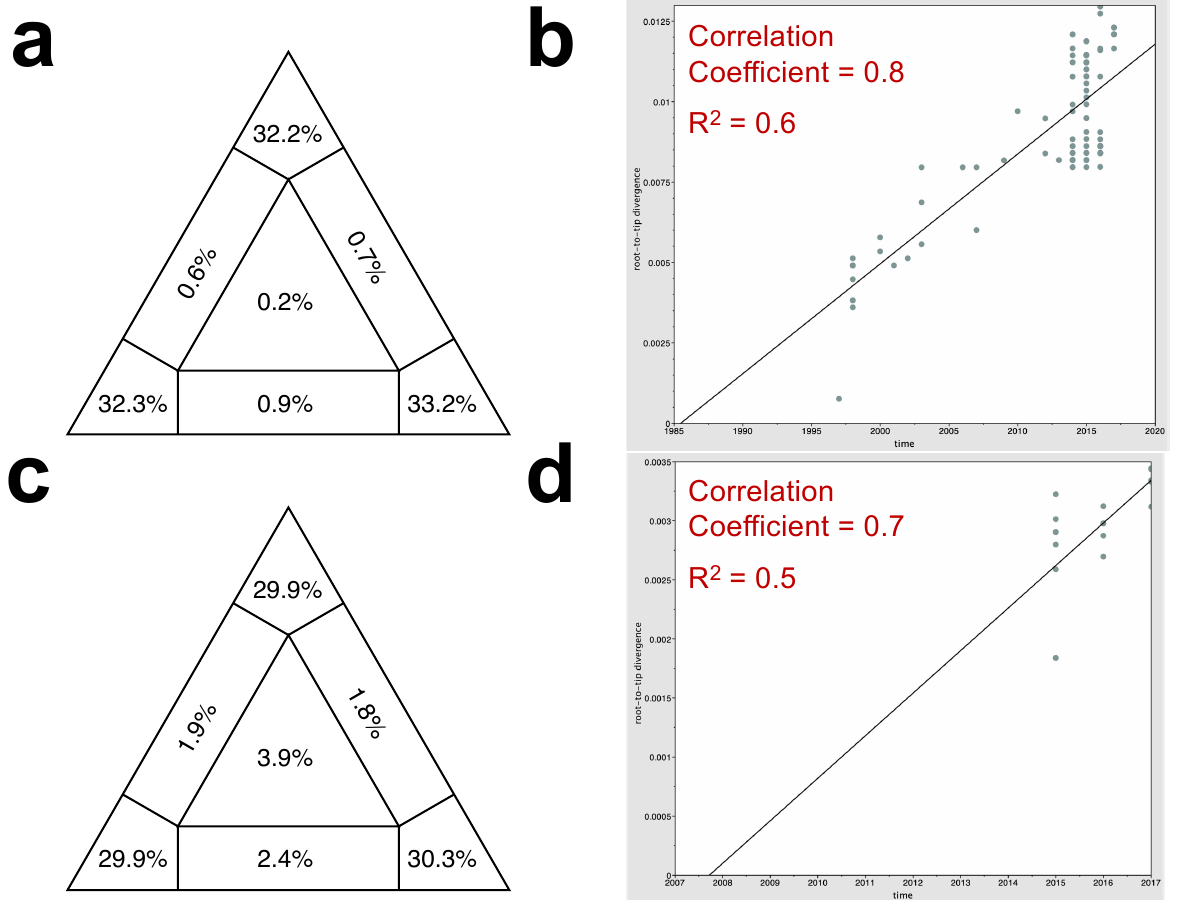

**Fig. S1. Estimation of phylogenetic and temporal signal from the DRC phylogenies.** Presence of phylogenetic signal in the dataset of the *V. cholerae* (a) dataset displayed figure 1 ( isolates collected by our group and downloaded from NCBI) , as well as (c) the dataset displayed figure 3 (isolates collected by our group only), was evaluated by likelihood mapping checking for alternative topologies (tips), unresolved quartets (center) and partly resolved quartets (edges) for each data set. (B) Linear regression of root-to-tip genetic distance within the ML phylogeny (tree in Figure 1) against sampling time for each taxon. Temporal resolution for (b) dataset displayed figure 1 or (d) dataset displayed figure 3 was assessed using the slope of the regression, with positive slope indicating sufficient temporal signal. Correlation coefficient, r, and R squared (R^2^) are reported in the temporal signal plot.

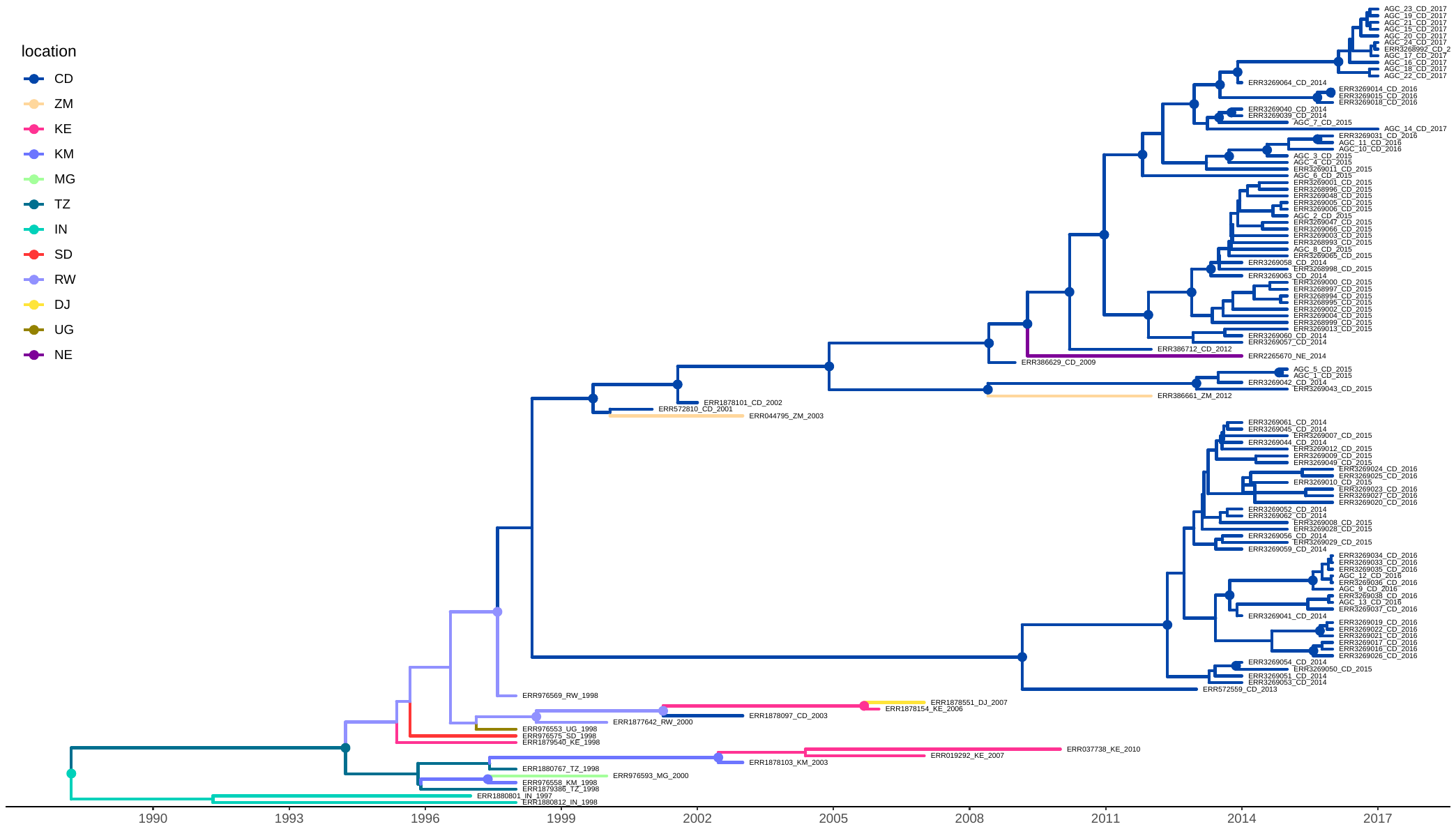

**Fig. S2. MCC in figure 1 tree with tips.** Phylogeny reported in figure 1 with tips. Branches of the phylogeny are scaled in time and colored by country of origin as shown in the legend (location). Circles in internal node indicate posterior probability (PP) support greater than 0.9 and the color indicate the ancestral country inferred by Bayesian phylogeography reconstruction.

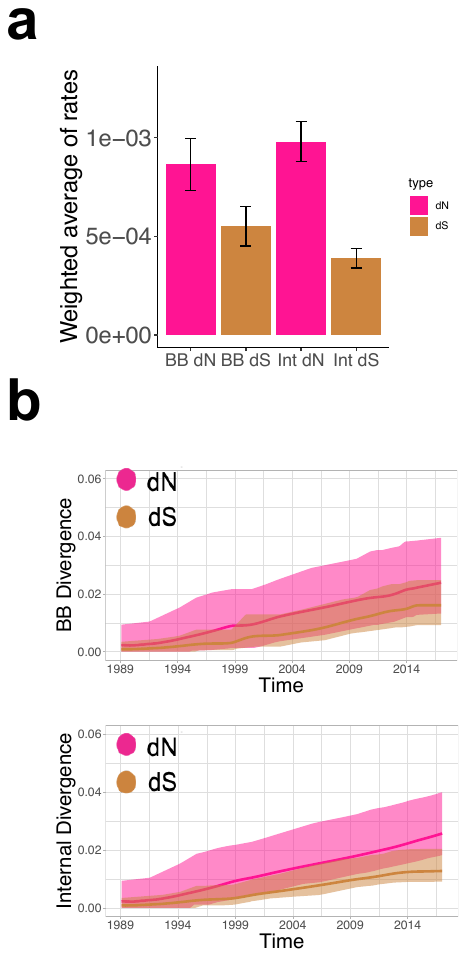

**Fig. S3. Tree, synonymous and nonsynonymous substitution rates.** (**a**) Weighted average of synonymous (dS) and nonsynonymous substitution (dN) rates of backbone (BB) and internal branches based on the Bayesian phylogeography tree in Fig. 1. (**b**) Absolute synonymous (tan) and non-synonymous (pink) divergence rates (y-axis) against time in years (x-axis) of backbone (BB) (top) and internal branches (bottom) of the phylogeny.

**Supplementary Tables**

**Table S1.** List of toxigenic *V. cholerae* O1 strains isolated from DRC in the present study.

| Strain | Isolation date | Health zone/Location | Serotype | Phage susceptibility |
| --- | --- | --- | --- | --- |
| AGC_1_CD_2015 | 4/30/2015 | North Kivu/ Kirotshe | Inaba | S |
| AGC_2_CD_2015 | 5/18/2015 | Goma/Buhimba | Inaba | S |
| AGC_3_CD_2015 | 5/20/2015 | Mutwanga | Inaba | R |
| AGC_4_CD_2015 | 3/7/2015 | Goma/Buhimba | Inaba | R |
| AGC_5_CD_2015 | 3/20/2015 | Goma/Buhimba | Inaba | S |
| AGC_6_CD_2015 | 7/26/2015 | Goma/Buhimba | Inaba | R |
| AGC_7_CD_2015 | 8/6/2015 | Goma/Buhimba | Inaba | S |
| AGC_8_CD_2015 | 8/6/2015 | Goma/Buhimba | Inaba | S |
| AGC_9_CD_2016 | 6/20/2016 | Maniema/ Kabambare | Ogawa | S |
| AGC_10_CD_2016 | 8/9/2016 | Karisimbi/ Hop Millitaire | Inaba | R |
| AGC_11_CD_2016 | 5/28/2016 | Alimbongo | Inaba | R |
| AGC_12_CD_2016 | 7/27/2016 | South Kivu/Fizi | Ogawa | S |
| AGC_13_CD_2016 | 8/8/2016 | South Kivu/Kimbilulenge | Ogawa | S |
| AGC_14_CD_2017 | 5/18/2017 | Kirotshe/Rubaya | Inaba | S |
| AGC_15_CD_2017 | 5/31/2017 | Rutshuru/Hgr | Inaba | S |
| AGC_16_CD_2017 | 6/10/2017 | Rutshuru/Hgr | Inaba | S |
| AGC_17_CD_2017 | 7/1/2017 | Nyiragongo/Turunga | Inaba | S |
| AGC_18__CD_2017 | 7/3/2017 | Goma/Hop.Provincial | Inaba | S^a^ |
| AGC_19_CD_2017 | 7/3/2017 | Goma/Hop.Provincial | Inaba | S |
| AGC_20_CD_2017 | 7/3/2019 | Goma/Hop.Provincial | Inaba | S |
| AGC_21_CD_2017 | 7/6/2017 | Karisimbi/Prison centrale | Inaba | S |
| AGC_22_CD_2017 | 7/14/2017 | Karisimbi/Majengo | Inaba | S^a^ |
| AGC_23_CD_2017 | 7/19/2017 | Karisimbi/Majengo | Inaba | R |
| AGC_24_CD_2017 | 07/15/2017 | Karisimbi/Majengo | Inaba | S^b^ |

* Susceptibility to a virulent ICP1 phage (ICP1_2017_A_DRC) to each of the *V. cholerae* strain is shown with strain yielding either complete resistance or form turbid plaque in response to phage infection as assayed by plaque assay.

**Table S2**. Complete list of all *V. cholerae* O1 strains and IPC1 phages used in the study. See Excel file.

**Table S3**. MLST analysis of the *V. cholerae* O1 strains isolated from DRC

| **Sample Name** | **MLST profile (ST)** |
| --- | --- |
| AGC_12_CD_2016 | 69 |
| AGC_13_CD_2016 | 69 |
| AGC_9_CD_2016 | 69 |
| AGC_1_CD_2015 | 515 |
| AGC_10_CD_2016 | 515 |
| AGC_11_CD_2016 | 515 |
| AGC_14_CD_2017 | 515 |
| AGC_15_CD_2017 | 515 |
| AGC_16_CD_2017 | 515 |
| AGC_17_CD_2017 | 515 |
| AGC_18_CD_2017 | 515 |
| AGC_19_CD_2017 | 515 |
| AGC_2_CD_2015 | 515 |
| AGC_20_CD_2017 | 515 |
| AGC_21_CD_2017 | 515 |
| AGC_22_CD_2017 | 515 |
| AGC_23_CD_2017 | 515 |
| AGC_24_CD_2017 | 515 |
| AGC_3_CD_2015 | 515 |
| AGC_4_CD_2015 | 515 |
| AGC_5_CD_2015 | 515 |
| AGC_6_CD_2015 | 515 |
| AGC_7_CD_2015 | 515 |
| AGC_8_CD_2015 | 515 |

**Table S4.** List of cholera patients’ stool samples obtained in DRC. Each stool sample was screened to isolate a unique ICP1 (ICP1_2017_A_DRC) phage using standard plaque assay All samples yielded toxigenic *V. cholerae* serogroup O1 strains with Inaba serotype.

| Sample ID | Origin of sample/ Location | Isolation time | Sensitivity of ICP1 (ICP1_2017_A_DRC) phage to *V. cholerae* O1 Inaba strain (AGC-15) |
| --- | --- | --- | --- |
| 1 | Vuhovi/Cs-kibwe | 10/8/2016 | R |
| 2 | Vuhovi/ Cs-kibwe | 10/8/2016 | R |
| 10 | Kirotshe/Sake | 3/11/2016 | R |
| 17 | Rutshuru/Cs-umoja | 11/17/2016 | R |
| 20 | Masisi/Cs-katale | 12/8/2016 | R |
| 23 | Kirotshe/Hgr | 26/12/2016 | R |
| 29 | Rutshuru/Hgr | 1/11/2017 | R |
| 31 | Rutshuru/Hgr | 1/11/2017 | R |
| 32 | Rutshuru/Hgr | 1/11/2017 | S |
| 33 | Rutshuru/Hgr | 1/11/2017 | R |
| 34 | Rutshuru/Hgr | 1/11/2017 | R |
| 36 | Masisi/Kitchanga | 2/15/2017 | R |
| 38 | Masisi/Kitchanga | 2/15/2017 | R |
| 39 | Kirotshe/Ngungu | 3/3/2017 | R |
| 48 | Goma/Hgr | 3/18/2017 | S |
| 49 | Nriragongo/Kibumba | 3/28/2017 | R |
| 55 | Rutshuru/Tongo | 3/30/2017 | S |
| 57 | Rutshuru/Tongo | 3/30/2017 | R |
| 60 | Pinga/Hgr | 4/3/2017 | S |
| 61 | Pinga/Hgr | 4/3/2017 | R |
| 65 | Pinga/Cs-riva | 4/3/2017 | R |
| 66 | Pinga/Hgr | 4/3/2017 | R |
| 68 | Kirotshe/Sake | 3/2/2017 | S |
| 71 | Kirotshe/Sake | 3/2/2017 | S |
| 74 | Kirotshe/Sake | 3/2/2017 | S |
| 77 | Nriragongo/Kiziba | 4/10/2017 | R |
| 80 | Rutshuru/Ntamugenga | 4/11/2017 | S |
| 81 | Rutshuru/Ntamugenga | 4/11/2017 | S |
| 82 | Rutshuru/Ntamugenga | 4/11/2017 | S |
| 85 | Kirotshe/Bweremana | 4/13/2017 | S |
| 87 | Nyiragongo/Kibumba | 4/15/2017 | S |
| 88 | Nyiragongo/Kibumba | 4/15/2017 | R |
| 89 | Nyiragongo/Kibumba | 4/15/2017 | R |
| 93 | Kirotshe/Mubambiro | 4/17/2017 | S |
| 94 | Kirotshe/Mubambiro | 4/17/2017 | S |
| 96 | Goma/Hgr | 4/17/2017 | R |
| 97 | Goma/Hgr | 4/17/2017 | S |
| 99 | Rutshuru/Hgr | 4/20/2017 | R |
| 105 | Goma/Hgr | 4/22/2017 | R |
| 106 | Goma/Hgr | 4/22/2017 | S |
| 107 | Kirotshe/Sake | 4/20/2017 | S |

**Table S5. List of mutation in phage resistant DRC *V. cholerae* strains detected in the LPS regions (VC0240-VC0269) of chromosome I, required for the biosynthesis of O-antigen.**

| Strain | ORFs^a^ | Predicted protein/functions | Position in the chromosome | Base change | Amino acid change due to codon change | Type of mutation | Phage sensitivity |
| --- | --- | --- | --- | --- | --- | --- | --- |
| AGC_3_CD_2015 | *VC0243* | GDP‑mannose 4,6‑dehydratase | 4532 | A→C | T127P (ACT→CCT) | Missense | R |
| AGC_4_CD_2015 | *VC0251* | acyl protein synthase/  acyl‑CoA reductase RfbN | 13638 | T→C | S506P (TCT→CCT) | Missense | R |
| AGC_6_CD_2015 | *VC0259* | lipopolysaccharide biosynthesis protein RfbV | 19921 | G→A | P133S (CCA→TCA) | Missense | R |
| AGC_10_CD_2016 | *VC0243* | GDP‑mannose 4,6‑dehydratase | 4532 | A→C | T127P (ACT→CCT) | Missense | R |
| AGC_11_CD_2016 | *VC0243* | GDP‑mannose 4,6‑dehydratase | 4532 | A→C | T127P (ACT→CCT) | Missense | R |
| AGC_23_CD_2017 | *VC0242* | Phosphomannomutase | 3301 | G→A | G181D (GGT→GAT) | Missense | R |
| AGC_18_CD_2017 | *VC0269* | mannose‑6‑phosphate isomerase | 29944 | Δ1 bp | coding (605/1200 nt) | Frameshift | S^b^ |
| AGC_22_CD_2017 | *VC0269* | mannose‑6‑phosphate isomerase | 29551 | Δ1 bp | coding (212/1200 nt) | Frameshift | S^b^ |
| AGC_24_CD_2017 | *VC0260* | mannosyl‑transferase | 21943 | Δ18 bp | coding (111‑128/1866 nt) | Inframe deletion | S^b^ |

^a^  Mutations are reported by comparison to *V. cholerae* O1 El Tor N16961 strain as reference.

^b^ Despite sustained non-synonymous mutation in O-antigen biosynthetic genes, DRC *V. cholerae* strains remained susceptible (either formed turbid plaque [AGC_18__CD_2017 and AGC_22_CD_2017] or typical plaque [AGC_24_CD_2017) to the virulent ICP1 phage infection (see the text for potential explanation of such event)
