## supplementary table S2 for "*Vibrio cholerae* multifaceted adaptive strategies in response to bacteriophage predation in an endemic region of the Democratic Republic of the Congo"

| *V. cholera* name | Collection Date | geographic location (country and/or sea) | geographic location (region and locality) | Lineage | Latitude | Longitude | Location in Map Fig 1# |
|---|---|---|---|---|---|---|---|
| AGC_1_CD_2015 | 2015 | Democratic Republic of the Congo | North Kivu/ Kirotshe | ST515 | -1.613051 | 29.03132 | 6 |
| AGC_10_CD_2016 | 2016 | Democratic Republic of the Congo | Karisimbi/ Hop Millitaire | ST515 | 1.5064 | 29.4508 | 10 |
| AGC_11_CD_2016 | 2016 | Democratic Republic of the Congo | Alimbongo | ST515 | -0.36879 | 29.156179 | 11 |
| AGC_12_CD_2016 | 2016 | Democratic Republic of the Congo | South Kivu/Fizi | ST515 | -4.30058 | 28.94212 | 12 |
| AGC_13_CD_2016 | 2016 | Democratic Republic of the Congo | South Kivu/Kimbilulenge | ST515 | -3.21838 | 28.25855 | 13 |
| AGC_14_CD_2017 | 2017 | Democratic Republic of the Congo | Kirotshe/Rubaya | ST515 | -1.546277 | 28.873122 | 14 |
| AGC_15_CD_2017 | 2017 | Democratic Republic of the Congo | Rutshuru/Hgr | ST515 | -1.188054595 | 29.4459123 | 15 |
| AGC_16_CD_2017 | 2017 | Democratic Republic of the Congo | Rutshuru/Hgr | ST515 | -1.188054595 | 29.4459123 | 15 |
| AGC_17_CD_2017 | 2017 | Democratic Republic of the Congo | Nyiragongo/Turunga | ST515 | -1.3527161 | 29.37873 | 16 |
| AGC_18_CD_2017 | 2017 | Democratic Republic of the Congo | Goma/Hop.Provincial | ST515 | -1.678865426 | 29.8 | 17 |
| AGC_19_CD_2017 | 2017 | Democratic Republic of the Congo | Goma/Hop.Provincial | ST515 | -1.678865426 | 29.8 | 17 |
| AGC_2_CD_2015 | 2015 | Democratic Republic of the Congo | Goma/Buhimba | ST69 | -1.621214 | 29.156623 | 7 |
| AGC_20_CD_2017 | 2017 | Democratic Republic of the Congo | Goma/Hop.Provincial | ST515 | -1.678865426 | 29.8 | 17 |
| AGC_21_CD_2017 | 2017 | Democratic Republic of the Congo | Karisimbi/Prison centrale | ST69 | -1.9 | 29 | 18 |
| AGC_22_CD_2017 | 2017 | Democratic Republic of the Congo | Karisimbi/Majengo | ST69 | -1.65388 | 29.5 | 19 |

| | | | | | | | |
|---|---|---|---|---|---|---|---|
| AGC_23_CD_2017 | 2017 | Democratic Republic of the Congo | Karisimbi/Majengo | ST515 | -1.65388 | 29.5 | 19 |
| AGC_24_CD_2017 | 2017 | Democratic Republic of the Congo | Karisimbi/Majengo | ST515 | -1.65388 | 29.5 | 19 |
| AGC_3_CD_2015 | 2015 | Democratic Republic of the Congo | Mutwanga | ST515 | 0.514939 | 25.191932 | 8 |
| AGC_4_CD_2015 | 2015 | Democratic Republic of the Congo | Goma/Buhimba | ST515 | -1.621214 | 29.156623 | 7 |
| AGC_5_CD_2015 | 2015 | Democratic Republic of the Congo | Goma/Buhimba | ST515 | -1.621214 | 29.156623 | 7 |
| AGC_6_CD_2015 | 2015 | Democratic Republic of the Congo | Goma/Buhimba | ST515 | -1.621214 | 29.156623 | 7 |
| AGC_7_CD_2015 | 2015 | Democratic Republic of the Congo | Goma/Buhimba | ST515 | -1.621214 | 29.156623 | 7 |
| AGC_8_CD_2015 | 2015 | Democratic Republic of the Congo | Goma/Buhimba | ST515 | -1.621214 | 29.156623 | 7 |
| AGC_9_CD_2016 | 2016 | Democratic Republic of the Congo | Maniema/ Kabambare | ST515 | -4.400901 | 27.765835 | 9 |
| ERR019292_KE_2007 | 2007 | Kenya | | | | | |
| ERR037738_KE_2010 | 2010 | Kenya | | | | | |
| ERR044795_ZM_2003 | 2003 | Zambia | | | | | |
| ERR1877642_RW_2000 | 2000 | Rwanda | | | | | |
| ERR1878097_CD_2003 | 2003 | Democratic Republic of Congo | | | | | |
| ERR1878101_CD_2002 | 2002 | Democratic Republic of Congo | Congo/Zaire | | -11.64112 | 27.51818 | 3 |
| ERR1878103_KM_2003 | 2003 | Comoros | | | | | |
| ERR1878154_KE_2006 | 2006 | Kenya | | | | | |
| ERR1878551_DJ_2007 | 2007 | Djibouti | | | | | |
| ERR1879386_TZ_1998 | 1998 | Tanzania | | | | | |
| ERR1879540_KE_1998 | 1998 | Kenya | | | | | |
| ERR1880767_TZ_1998 | 1998 | Tanzania | | | | | |

| | | | | | | |
|---|---|---|---|---|---|---|
| ERR1880801_IN_1997 | 1997 | India | | | | |
| ERR1880812_IN_1998 | 1998 | India | | | | |
| ERR2265670_NE_2014 | 2014 | Niger | | | | |
| ERR3268992_CD_2017 | 2017 | Democratic Republic of the Congo | Minova | -4.32153 | 15.31185 | 3 |
| ERR3268993_CD_2015 | 2015 | Democratic Republic of the Congo | Goma | -1.6835 | 29.2356 | 17 |
| ERR3268994_CD_2015 | 2015 | Democratic Republic of the Congo | Goma | -1.6835 | 29.2356 | 17 |
| ERR3268995_CD_2015 | 2015 | Democratic Republic of the Congo | Goma | -1.6835 | 29.2356 | 17 |
| ERR3268996_CD_2015 | 2015 | Democratic Republic of the Congo | Goma | -1.6835 | 29.2356 | 17 |
| ERR3268997_CD_2015 | 2015 | Democratic Republic of the Congo | Goma | -1.6835 | 29.2356 | 17 |
| ERR3268998_CD_2015 | 2015 | Democratic Republic of the Congo | Goma | -1.6835 | 29.2356 | 17 |
| ERR3268999_CD_2015 | 2015 | Democratic Republic of the Congo | Goma | -1.6835 | 29.2356 | 17 |
| ERR3269000_CD_2015 | 2015 | Democratic Republic of the Congo | Goma | -1.6835 | 29.2356 | 17 |
| ERR3269001_CD_2015 | 2015 | Democratic Republic of the Congo | Goma | -1.6835 | 29.2356 | 17 |
| ERR3269002_CD_2015 | 2015 | Democratic Republic of the Congo | Goma | -1.6835 | 29.2356 | 17 |
| ERR3269003_CD_2015 | 2015 | Democratic Republic of the Congo | Goma | -1.6835 | 29.2356 | 17 |
| ERR3269004_CD_2015 | 2015 | Democratic Republic of the Congo | Goma | -1.6835 | 29.2356 | 17 |
| ERR3269005_CD_2015 | 2015 | Democratic Republic of the Congo | Goma | -1.6835 | 29.2356 | 17 |

| | | | | | | | |
|---|---|---|---|---|---|---|---|
| ERR3269006_CD_2015 | 2015 | Democratic Republic of the Congo | Goma | ST515 | -1.6835 | 29.2356 | 17 |
| ERR3269007_CD_2015 | 2015 | Democratic Republic of the Congo | Fizi-Baraka | ST515 | -4.30058 | 28.94212 | 12 |
| ERR3269008_CD_2015 | 2015 | Democratic Republic of the Congo | Fizi-Baraka | ST515 | -4.30058 | 28.94212 | 12 |
| ERR3269009_CD_2015 | 2015 | Democratic Republic of the Congo | Fizi-Baraka | ST515 | -4.30058 | 28.94212 | 12 |
| ERR3269010_CD_2015 | 2015 | Democratic Republic of the Congo | Fizi-Baraka | ST515 | -4.30058 | 28.94212 | 12 |
| ERR3269011_CD_2015 | 2015 | Democratic Republic of the Congo | Bukavu | ST515 | -2.50316 | 28.85309 | 21 |
| ERR3269012_CD_2015 | 2015 | Democratic Republic of the Congo | Fizi-Baraka | ST515 | -4.30058 | 28.94212 | 12 |
| ERR3269013_CD_2015 | 2015 | Democratic Republic of the Congo | Minova | ST515 | -4.32153 | 15.31185 | 3 |
| ERR3269014_CD_2016 | 2016 | Democratic Republic of the Congo | Fizi-Baraka | ST515 | -4.30058 | 28.94212 | 12 |
| ERR3269015_CD_2016 | 2016 | Democratic Republic of the Congo | Fizi-Baraka | ST515 | -4.30058 | 28.94212 | 12 |
| ERR3269016_CD_2016 | 2016 | Democratic Republic of the Congo | Fizi-Baraka | ST515 | -4.30058 | 28.94212 | 12 |
| ERR3269017_CD_2016 | 2016 | Democratic Republic of the Congo | Fizi-Baraka | ST515 | -4.30058 | 28.94212 | 12 |
| ERR3269018_CD_2016 | 2016 | Democratic Republic of the Congo | Fizi-Baraka | ST515 | -4.30058 | 28.94212 | 12 |
| ERR3269019_CD_2016 | 2016 | Democratic Republic of the Congo | Fizi-Baraka | ST515 | -4.30058 | 28.94212 | 12 |
| ERR3269020_CD_2016 | 2016 | Democratic Republic of the Congo | Uvira | ST515 | -3.38413 | 29.1415 | 20 |
| ERR3269021_CD_2016 | 2016 | Democratic Republic of the Congo | Uvira | ST515 | -3.38413 | 29.1415 | 20 |

| | | | | | | | |
|---|---|---|---|---|---|---|---|
| ERR3269022_CD_2016 | 2016 | Democratic Republic of the Congo | Uvira | ST69 | -3.38413 | 29.1415 | 20 |
| ERR3269023_CD_2016 | 2016 | Democratic Republic of the Congo | Uvira | ST69 | -3.38413 | 29.1415 | 20 |
| ERR3269024_CD_2016 | 2016 | Democratic Republic of the Congo | Uvira | ST69 | -3.38413 | 29.1415 | 20 |
| ERR3269025_CD_2016 | 2016 | Democratic Republic of the Congo | Uvira | ST69 | -3.38413 | 29.1415 | 20 |
| ERR3269026_CD_2016 | 2016 | Democratic Republic of the Congo | Uvira | ST515 | -3.38413 | 29.1415 | 20 |
| ERR3269027_CD_2016 | 2016 | Democratic Republic of the Congo | Fizi-Baraka | ST69 | -4.30058 | 28.94212 | 12 |
| ERR3269028_CD_2015 | 2015 | Democratic Republic of the Congo | Fizi-Baraka | ST515 | -4.30058 | 28.94212 | 12 |
| ERR3269029_CD_2015 | 2015 | Democratic Republic of the Congo | Fizi-Baraka | ST515 | -4.30058 | 28.94212 | 12 |
| ERR3269031_CD_2016 | 2016 | Democratic Republic of the Congo | Alimbongo | ST515 | -0.3692 | 29.155569 | 8 |
| ERR3269033_CD_2016 | 2016 | Democratic Republic of the Congo | Kabambare | ST69 | -4.68967 | 27.69298 | 9 |
| ERR3269034_CD_2016 | 2016 | Democratic Republic of the Congo | Fizi-Baraka | ST69 | -4.30058 | 28.94212 | 12 |
| ERR3269035_CD_2016 | 2016 | Democratic Republic of the Congo | Fizi-Baraka | ST515 | -4.30058 | 28.94212 | 12 |
| ERR3269036_CD_2016 | 2016 | Democratic Republic of the Congo | Fizi-Baraka | ST69 | -4.30058 | 28.94212 | 12 |
| ERR3269037_CD_2016 | 2016 | Democratic Republic of the Congo | Kimbilulenge | ST69 | -4.32153 | 15.31185 | 3 |
| ERR3269038_CD_2016 | 2016 | Democratic Republic of the Congo | Kimbilulenge | ST69 | -4.32153 | 15.31185 | 3 |
| ERR3269039_CD_2014 | 2014 | Democratic Republic of the Congo | Fizi-Baraka | ST69 | -4.30058 | 28.94212 | 12 |

| | | | | | | | |
|---|---|---|---|---|---|---|---|
| ERR3269040_CD_2014 | 2014 | Democratic Republic of the Congo | Fizi-Baraka | ST69 | -4.30058 | 28.94212 | 12 |
| ERR3269041_CD_2014 | 2014 | Democratic Republic of the Congo | Fizi-Baraka | ST69 | -4.30058 | 28.94212 | 12 |
| ERR3269042_CD_2014 | 2014 | Democratic Republic of the Congo | Alimbongo | ST69 | -0.3692 | 29.155569 | 8 |
| ERR3269043_CD_2015 | 2015 | Democratic Republic of the Congo | Masisi | ST69 | -1.3527161 | 29.37873 | 16 |
| ERR3269044_CD_2014 | 2014 | Democratic Republic of the Congo | Fizi-Baraka | ST69 | -4.30058 | 28.94212 | 12 |
| ERR3269045_CD_2014 | 2014 | Democratic Republic of the Congo | Fizi-Baraka | ST69 | -4.30058 | 28.94212 | 12 |
| ERR3269047_CD_2015 | 2015 | Democratic Republic of the Congo | Goma | ST69 | -1.6835 | 29.2356 | 17 |
| ERR3269048_CD_2015 | 2015 | Democratic Republic of the Congo | Goma | ST515 | -1.6835 | 29.2356 | 17 |
| ERR3269049_CD_2015 | 2015 | Democratic Republic of the Congo | Goma | ST69 | -1.6835 | 29.2356 | 17 |
| ERR3269050_CD_2015 | 2015 | Democratic Republic of the Congo | Goma | ST69 | -1.6835 | 29.2356 | 17 |
| ERR3269051_CD_2014 | 2014 | Democratic Republic of the Congo | Uvira | ST69 | -3.38413 | 29.1415 | 20 |
| ERR3269052_CD_2014 | 2014 | Democratic Republic of the Congo | Uvira | ST69 | -3.38413 | 29.1415 | 20 |
| ERR3269053_CD_2014 | 2014 | Democratic Republic of the Congo | Uvira | ST69 | -3.38413 | 29.1415 | 20 |
| ERR3269054_CD_2014 | 2014 | Democratic Republic of the Congo | Uvira | ST69 | -3.38413 | 29.1415 | 20 |
| ERR3269056_CD_2014 | 2014 | Democratic Republic of the Congo | Uvira | ST515 | -3.38413 | 29.1415 | 20 |
| ERR3269057_CD_2014 | 2014 | Democratic Republic of the Congo | Fizi-Baraka | ST515 | -4.30058 | 28.94212 | 12 |

| | | | | | | | |
|---|---|---|---|---|---|---|---|
| ERR3269058_CD_2014 | 2014 | Democratic Republic of the Congo | Fizi-Baraka | ST69 | -4.30058 | 28.94212 | 12 |
| ERR3269059_CD_2014 | 2014 | Democratic Republic of the Congo | Goma | ST515 | -1.6835 | 29.2356 | 17 |
| ERR3269060_CD_2014 | 2014 | Democratic Republic of the Congo | Fizi-Baraka | ST515 | -4.30058 | 28.94212 | 12 |
| ERR3269061_CD_2014 | 2014 | Democratic Republic of the Congo | Fizi-Baraka | ST69 | -4.30058 | 28.94212 | 12 |
| ERR3269062_CD_2014 | 2014 | Democratic Republic of the Congo | Goma | ST69 | -1.6835 | 29.2356 | 17 |
| ERR3269063_CD_2014 | 2014 | Democratic Republic of the Congo | Fizi-Baraka | ST613 | -4.30058 | 28.94212 | 12 |
| ERR3269064_CD_2014 | 2014 | Democratic Republic of the Congo | Fizi-Baraka | ST612 | -4.30058 | 28.94212 | 12 |
| ERR3269065_CD_2015 | 2015 | Democratic Republic of the Congo | Goma | ST515 | -1.6835 | 29.2356 | 17 |
| ERR3269066_CD_2015 | 2015 | Democratic Republic of the Congo | Goma | ST515 | -1.6835 | 29.2356 | 17 |
| ERR386629_CD_2009 | 2009 | Democratic Republic of Congo | | | -5.91312 | 29.20005 | 4 |
| ERR386661_ZM_2012 | 2012 | Zambia | | | | | |
| ERR386712_CD_2012 | 2012 | Democratic Republic of Congo | Biyela | ST515 | -4.32153 | 15.31185 | 5 |
| ERR572559_CD_2013 | 2013 | Democratic Republic of Congo | Nyemba | ST69 | -8.16966 | 25.38507 | 1 |
| ERR572810_CD_2001 | 2001 | Democratic Republic of Congo | Ankoro | | -6.75053 | 26.94274 | 2 |
| ERR976553_UG_1998 | 1998 | Uganda | | | | | |
| ERR976558_KM_1998 | 1998 | Comoros | | | | | |
| ERR976569_RW_1998 | 1998 | Rwanda | | | | | |
| ERR976575_SD_1998 | 1998 | Sudan | | | | | |
| ERR976593_MG_2000 | 2000 | Madagascar | | | | | |

| Phage name | Collection Date | geographic location (country and/or sea) | geographic location (region and locality) | Lineage | Latitude | Longitude | Location in Map Fig 2# |
|---|---|---|---|---|---|---|---|
| DRC32 | 1/11/2017 | Democratic Republic of Congo | Rutshuru/Hgr | | -1.188054595 | 29.4459123 | 22 |
| DRC48 | 3/18/2017 | Democratic Republic of Congo | Goma/Hgr | | -1.678865426 | 29.22441067 | 17 |
| DRC55 | 3/30/2017 | Democratic Republic of Congo | Rutshuru/Tongo | | -1.209704343 | 29.27433726 | 23 |
| DRC71 | 3/2/2017 | Democratic Republic of Congo | Kirotshe/Sake | | -1.571673852 | 29.0497398 | 24 |
| DRC74 | 3/2/2017 | Democratic Republic of Congo | Kirotshe/Sake | | -1.571673852 | 29.0497398 | 24 |
| DRC82 | 4/11/2017 | Democratic Republic of Congo | Rutshuru/Ntamugenga | | -1.194247498 | 29.44567655 | 15 |
| DRC87 | 4/15/2017 | Democratic Republic of Congo | Nyiragongo/Kibumba | | -2.999999865 | 28.46649529 | 25 |
| DRC106 | 4/22/2017 | Democratic Republic of Congo | Goma/Hgr | | -1.678865426 | 29.22441067 | 22 |
